# Citation Contamination of Systematic Review Literature in the Life Sciences by Paper Mills

**DOI:** 10.1101/2024.12.26.24319521

**Authors:** Gengyan Tang, Hao Cai

## Abstract

**Importance:** Paper mills are systematic fraud organizations that mass-produce fabricated papers, submit them under researchers’ names, and profit by charging fees or selling authorship. Their products have infiltrated scientific literature databases, yet the academic community remains uncertain about their potential impact on scholarly research.

**Objective:** Our study investigates the citation contamination of systematic reviews in the life sciences by paper mill articles, aiming to determine whether these fabricated papers undermine the status of systematic reviews as the “gold standard” of evidence synthesis.

**Evidence Review:** We conducted a cross-sectional study of 100,000 systematic reviews published in the life sciences between 2013 and 2023, as indexed in the Web of Science. We extracted their references and matched them against retracted articles in the Retraction Watch Database, specifically those retracted due to paper mills. Descriptive statistical analyses were used to characterize the features of contaminated systematic reviews, including their subject areas, journals of publication, and citation patterns.

**Findings:** A total of 179 systematic reviews were contaminated by paper mill articles, representing a contamination rate of 0.179%. Although the overall extent of contamination was small, an increasing trend was observed. Notably, 61 citations occurred after the articles had been retracted. Oncology was the most severely affected field. Four systematic reviews each cited five or more paper mill articles; all were published by journals under the same academic publisher. Among those citing three or more paper mill articles, 23 (76.67%) were published in journals ranked in the top 50% of their respective fields by impact factor (i.e., those classified as Q1 or Q2).

**Conclusions and Relevance:** Systematic reviews, as comprehensive syntheses of high-quality evidence, must not incorporate systematically fabricated articles. The scientific community should remain vigilant about the growing trend of paper mill contamination in life sciences systematic reviews. Correcting and retracting previously contaminated reviews, and developing new tools to help researchers identify potential paper mill articles in literature databases, will be essential steps to ensure the integrity of evidence synthesis.

**Key Points:** *Question:* Do paper mill articles contaminate systematic reviews in the life sciences, and what is their potential impact on the integrity of evidence synthesis?

*Findings:* In a cross-sectional study of 100,000 systematic reviews published between 2013 and 2023, 0.179% were contaminated by paper mill articles, with an increasing trend observed over time. Oncology was the most affected field, and 61 post-retraction citations were identified.

*Meaning:* These findings underscore the need for heightened vigilance, systematic correction of contaminated reviews, and development of tools to identify paper mill articles to safeguard the integrity of evidence synthesis.

## 1. Introduction

Systematic reviews are considered a method for synthesizing high-quality evidence in the life sciences, integrating data from different studies to generate new aggregated results or conclusions. They may also combine diverse types of evidence to explore or interpret significance, while identifying knowledge gaps to guide future research ^1^ ^2^ ^3^. Systematic reviews have become a cornerstone of modern medical practice and are often regarded as the “gold standard” of evidence ^4^. Academic journals in the life sciences increasingly emphasize the importance of systematic reviews.

Discussions on systematic reviews largely focus on potential biases, particularly in evidence selection. For instance, the preference of academic journals for positive results ^5^ ^6^ ^7^ may lead systematic reviews to similarly favor positive findings during evidence selection ^8^. The neglect of grey literature ^9^ ^10^ and biases caused by language restrictions ^11^ ^12^ can also result in significant distortions in the evidence base of systematic reviews. Although systematic reviews are considered objective and scientific processes, the inclusion of low-quality or biased evidence can render them misleading or even harmful, especially when their conclusions influence clinical decision-making ^13^. While the PRISMA 2020 statement provides a framework to help life science researchers mitigate the risk of selection bias ^14^, the rapidly evolving issue in research integrity—the emergence of paper mills producing large volumes of fabricated evidence— remains underaddressed.

Paper mills refer to organizations that profit by producing fake manuscripts and selling authorship positions to academic clients ^15^. In one report, researchers conducting a systematic review on stroke treatments discovered that many papers included suspicious data and questionable images, potentially endangering the integrity of systematic reviews as a method of evidence synthesis. These papers were suspected to be products of paper mills ^16^. Fake papers generated by paper mills have infiltrated scientific databases on a large scale ^17^ ^18^ ^19^, yet their impacts remain poorly understood. In November 2024, the Committee on Publication Ethics (COPE) released an expert consensus through the United2Act working group, specifically addressing the governance of paper mills. The consensus called for extensive research into the effects of paper mills on scientific research, as current evidence is primarily anecdotal ^20^.

Previous studies on paper mills have examined their potential characteristics ^21^ ^22^ ^23^, identifying that, unlike ordinary retracted papers (e.g., those retracted for self-correction), papers produced by paper mills represent large-scale, systematic fabrication. These fabrications include manipulated images ^24^, falsified data ^25^, and citation manipulation ^17^. They are often published in journals with relatively high impact factors ^18^. In this study, we provide further evidence to demonstrate whether papers produced by paper mills have contaminated systematic review literature in the life sciences. Specifically, we examined the references cited in systematic reviews within this field, identified contaminated papers, and analyzed their potential characteristics. This research contributes to a deeper understanding of paper mills in the field of research integrity and offers insights to help safeguard systematic reviews in the life sciences from contamination by paper mill products.

## 2. Materials and Methods

### 2.1. Data Management and Analysis Tools

Data management, categorization, and filtering were conducted using Microsoft Excel (Version 2403 Build 16.0.17425.20176, 2021, Microsoft Corporation, Redmond, Washington, USA, https://office.microsoft.com/excel). For citation matching, data analysis, and result visualization, PyCharm 2024.1.7 (JetBrains, 2024, Prague, Czech Republic, https://www.jetbrains.com/pycharm/) was employed.

### 2.2. Data Acquisition and Filtering

Data were obtained from three primary sources:

1. Retraction Watch Database. The first dataset was sourced from the Retraction Watch Database via the Center for Scientific Integrity on December 6, 2024 (https://gitlab.com/crossref/retraction-watch-data). This dataset comprised bibliographic information on 58,736 retracted publications ^26^, including original publication dates, retraction dates, DOIs, and reasons for retraction. For this study, we filtered entries where the retraction reason included the term “Paper Mill,” resulting in a dataset of 7,887 retracted publications for analysis.
2. Web of Science (WoS). The second dataset was retrieved from the Science Citation Index-Expanded (SCI-Expanded) within the Web of Science (WoS) database, known for its high indexing standards, comprehensive citation links, and detailed metadata ^27^. Using the keyword “Systematic Review,” we identified 343,080 publications (as of December 6, 2024). Inclusion criteria were applied as follows: (1) Publications dated between 2013 and 2023. (2) Document types classified as “Review” or “Research Article.” (3) Research areas categorized under “Life Sciences & Biomedicine” (https://webofscience.help.clarivate.com/Content/research-areas.html?Highlight=research%20area). After applying these criteria, 177,070 publications were identified. To manage the volume, we extracted the first 100,000 systematic review publications for further analysis. This dataset included details such as publication dates, journal names, DOIs, authors, and the DOIs of cited references. The full search strategy can be accessed here: https://www.webofscience.com/wos/woscc/summary/0377883e-8288-47da-9d07-826200725525-013320eca2/relevance/1.
3. Journal Citation Reports (JCR). The third dataset was obtained from Clarivate’s JCR and provided information on the impact factor quartiles of journals publishing systematic reviews that cited retracted publications, specifically those retracted due to paper mill activities.

### 2.3. Citation Matching

A Python script (details in Supplementary Material 1) was developed to extract key bibliographic information from the WoS dataset, including article titles, authors, journals, publication years, and references. From the references, all cited DOIs were extracted and matched against the DOIs in the Retraction Watch Database. This process identified WoS articles that cited retracted publications. The matched records included metadata such as the original publication date and retraction date of the cited papers, supporting an analysis of the temporal relationship between citation and retraction. For each matched DOI, detailed information on the citing article (e.g., title, authors, journal) and the corresponding retracted article was extracted. The final results were compiled into a structured dataset and saved as a CSV file for subsequent analysis.

### 2.4. Data Processing and Analysis

The analysis was performed in five dimensions:

1. Contamination overview. Using descriptive statistical analysis, we calculated the contamination rate of systematic reviews affected by papers produced by paper mills, as well as the distribution of citation counts for these contaminated papers.
2. Temporal analysis. (1) Time lag: The difference between the publication date of systematic review articles and the retraction date of cited articles, reflecting whether authors were diligent in checking citation validity. (2) Temporal trends: The annual number of systematic review articles identified as citing retracted articles and the frequency of citations to paper mill articles.
3. Disciplinary analysis. The disciplinary distribution of systematic review articles was examined to identify potential clustering in specific research areas.
4. Citation patterns. Systematic review articles citing three or more retracted articles were analyzed to reveal citation behaviors and patterns.
5. Journal distribution. The journals publishing systematic reviews citing retracted articles were analyzed to identify whether specific journals disproportionately hosted such publications.

This multifaceted approach sheds light on the influence of paper mills on systematic reviews in the life sciences.

## 3. Results

### 3.1. Contamination Rate

As shown in Figure 1, a total of 179 systematic reviews in the life sciences domain were identified as contaminated, resulting in a contamination rate of 0.179%. They cited papers from paper mills a total of 246 times. Among these contaminated reviews, 149 (83.24%) cited at least one paper originating from a paper mill. Additionally, 30 reviews (16.76%) cited two or more papers produced by paper mills. Notably, two reviews cited six papers from paper mills, and one review cited as many as 13 papers.

**Fig. 1.**
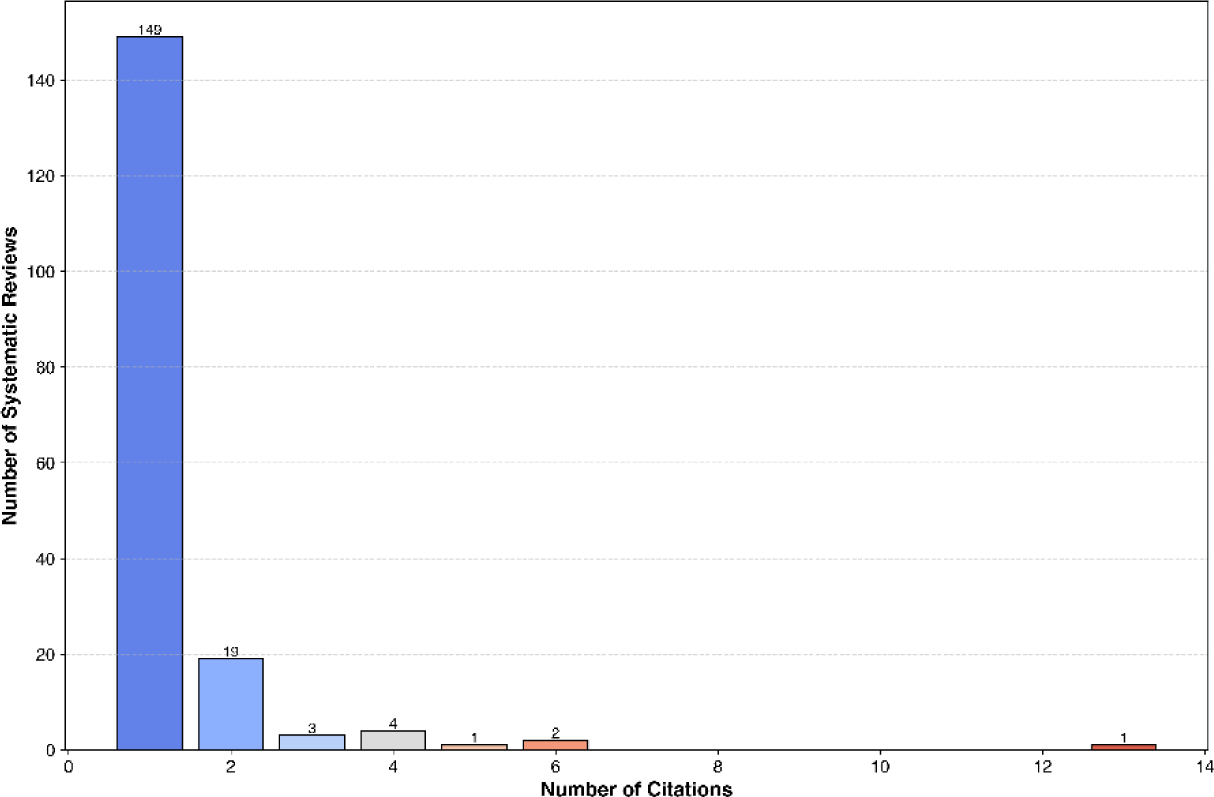
contamination of systematic reviews by paper mill articles

### 3.2. Temporal Differences and Distribution

Analysis of temporal differences (Figure 2a) revealed that the majority of citations occurred before the retraction of the paper mill articles, with a particular concentration within 500 days prior to retraction. However, 61 citations occurred after the retraction of the paper mill articles, among which 10 citations were made more than 500 days post-retraction. The mean time difference between citations and retractions was 455.2 days prior to retraction.

**Fig. 2.**
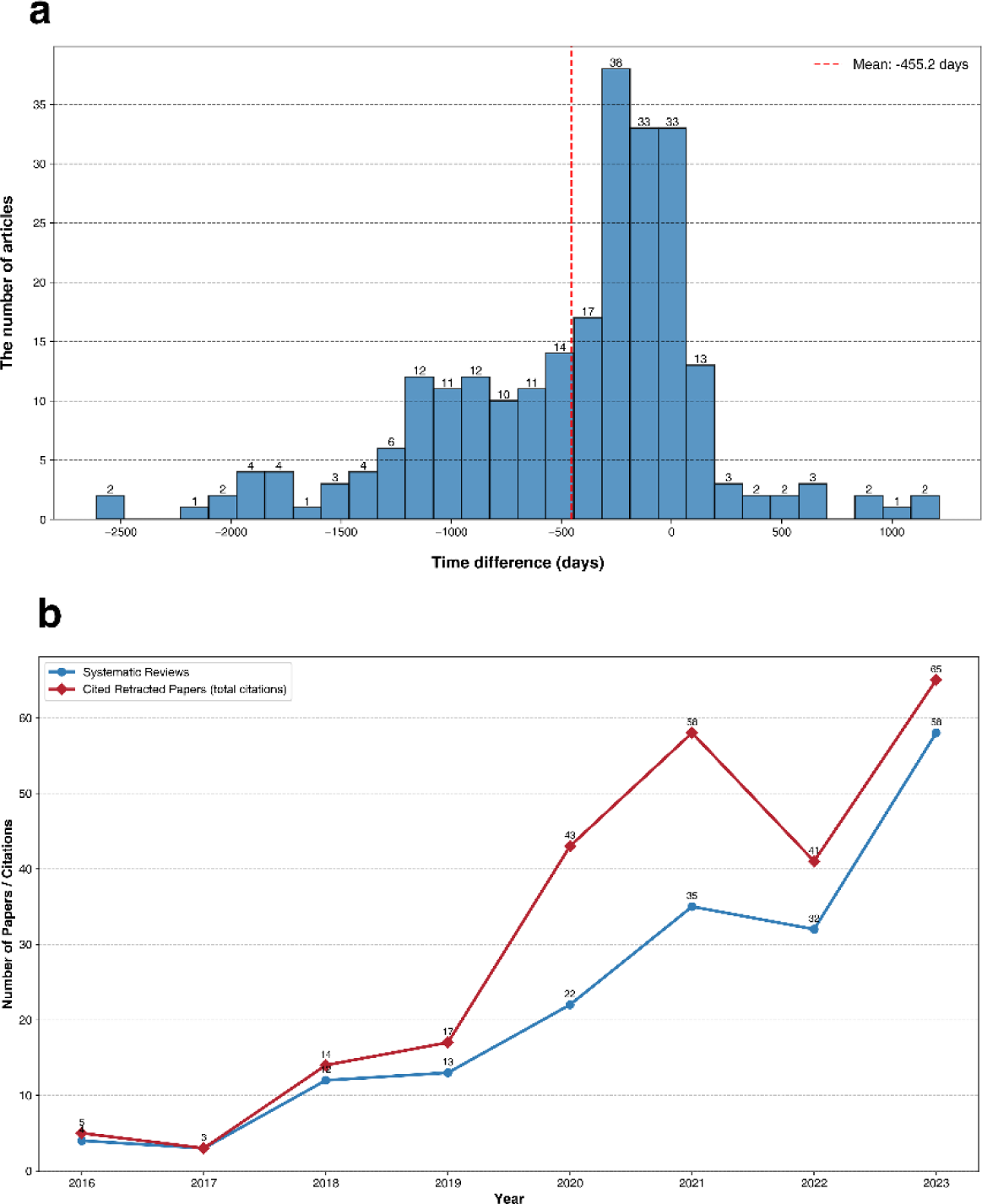
(a) temporal distribution of citations relative to retraction; (b) annual trends in systematic reviews citing paper mill articles

Figure 2b illustrates an increasing trend in the number of systematic reviews in the life sciences citing paper mill articles. No systematic reviews citing paper mill articles were identified in 2013, 2014, or 2015. Starting in 2016 (*n* = 4), the number of systematic reviews citing paper mill articles began to increase, reaching 58 in 2023—an increase of 1350%. The number of citations rose from 5 in 2016 to 65 in 2023, representing a 1200% increase.

### 3.3. Research Areas

As shown in Figure 3, systematic reviews in oncology represent the most heavily contaminated research area, with 28 systematic reviews citing articles from paper mills. This is followed by biochemistry & molecular biology (*n* = 24), general & internal medicine (*n* = 18), cell biology (*n* = 11), pharmacology &pharmacy (*n* = 10), and environmental sciences & ecology (*n* = 10).

**Fig. 3.**
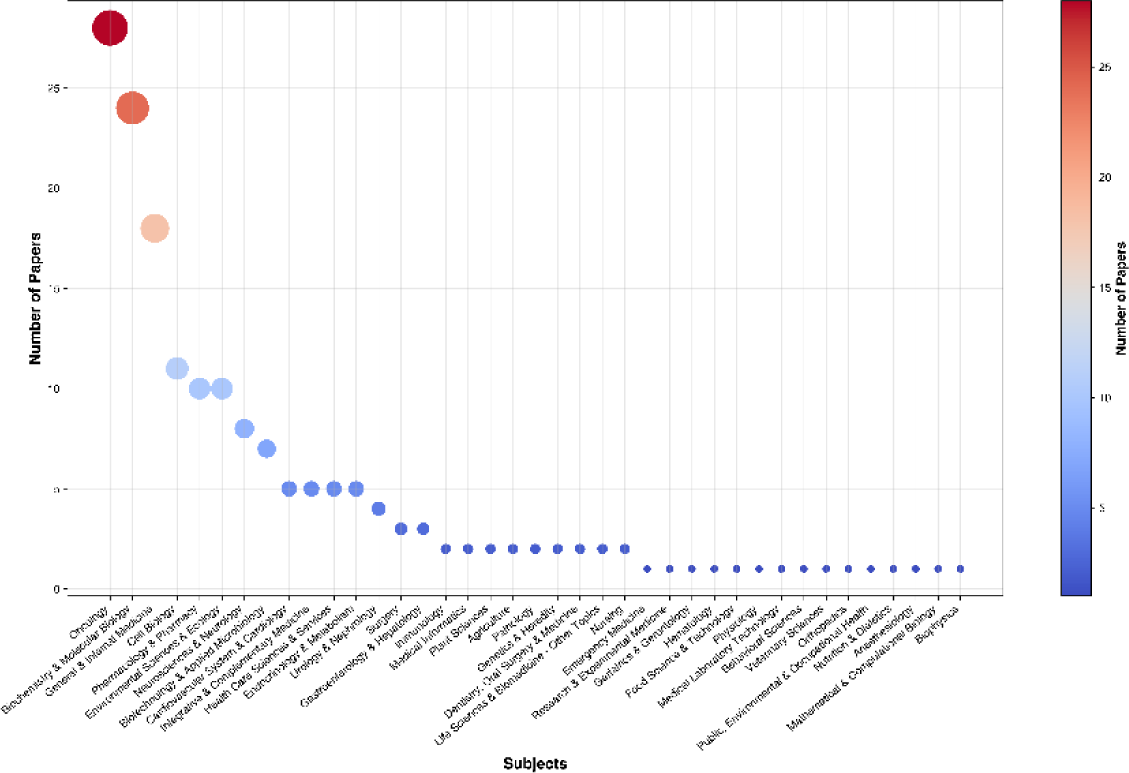
research areas affected by paper mill contamination in systematic reviews

Research areas with fewer than 10 but at least 5 contaminated systematic reviews include neurosciences & neurology (*n* = 8), biotechnology & applied microbiology (*n* = 7), cardiovascular system & cardiology (*n* = 5), integrative & complementary medicine (*n* = 5), health care sciences & services (*n* = 5), and endocrinology & metabolism (*n* = 5).

### 3.4. Citation Patterns

We examined systematic review articles that cited paper mill articles three or more times to identify heavily contaminated systematic reviews. The corresponding article IDs, titles, and DOIs are provided in the Supplementary Material 2. As shown in Figure 4, among 11 systematic reviews, four articles ^28^ ^29^ ^30^ ^31^ cited paper mill articles five or more times. All four reviews were published in academic journals under the MDPI umbrella. Notably, article ^29^ cited paper mill articles 13 times. A detailed review of this article revealed that all 13 citations occurred in the results section. Articles ^28^ ^30^ ^31^ exhibited a similar citation pattern.

**Fig. 4.**
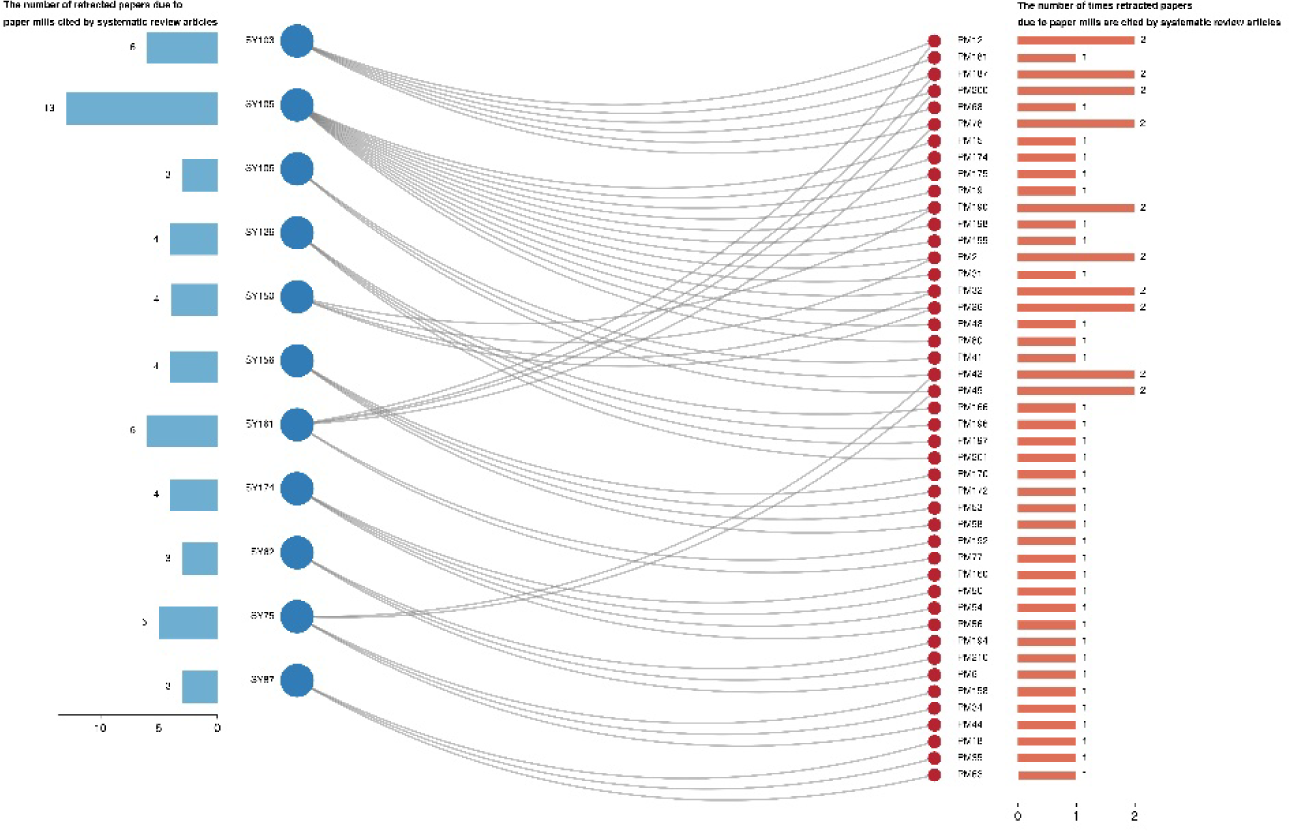
heavily contaminated systematic reviews and their citation patterns

According to Dimensions data, article ^29^ has been cited by 19 other publications, article ^30^ by 22, article ^31^ by 16, and article ^28^ by 20. Furthermore, article ^28^ was cited in a rule published by the U.S. federal government ^32^. This rule, issued on January 18, 2023, added all types of uterine cancer, including endometrial cancer, to the list of World Trade Center (WTC)-related health conditions.

### 3.5. Journal Distribution

As shown in Figure 5, we analyzed the journals that published systematic reviews citing paper mill articles. The central section of Figure 5 highlights the systematic reviews that cited paper mill articles at least twice (*n* = 30). Among these reviews, 18 (60%) were published in academic journals classified into the first quartile (Q1; top 25% by impact factor), 5 (16.67%) in the second quartile (Q2; 25%–50%), 1 (3.33%) in the third quartile (Q3; 50%–75%), and 4 (13.33%) in the fourth quartile (Q4; below 75%).

**Fig. 5.**
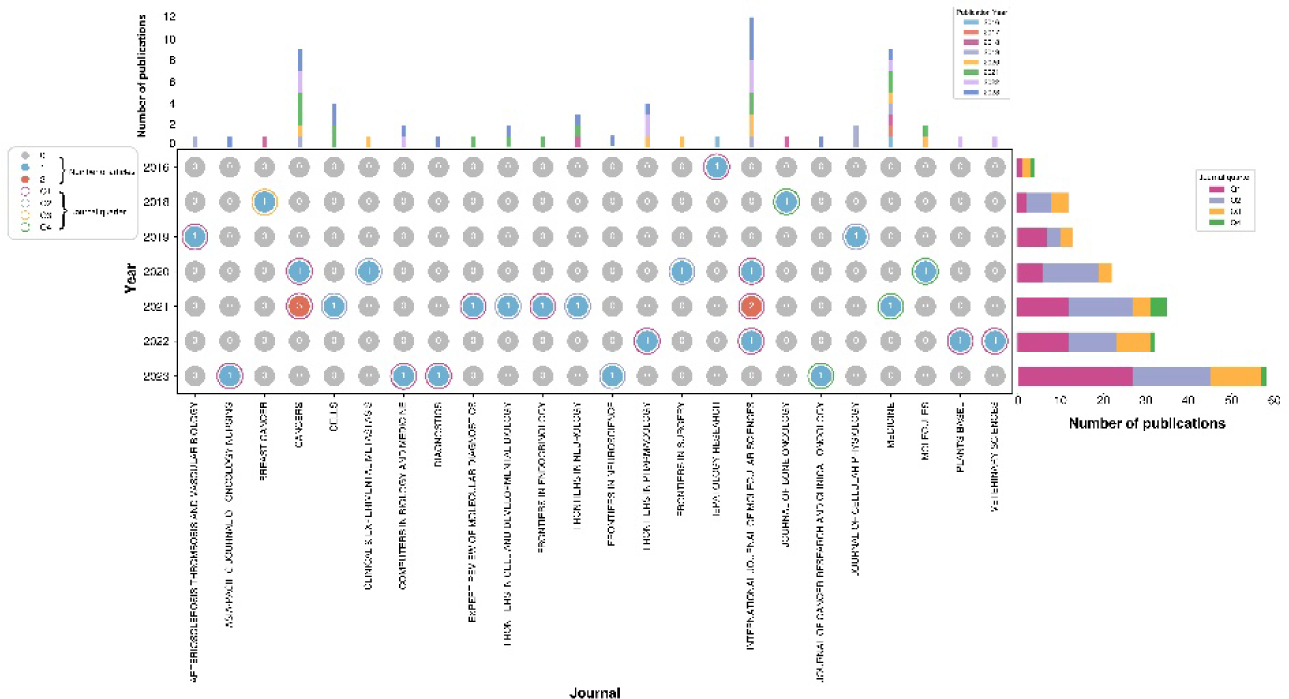
journal distribution and publication trends of systematic reviews citing paper mill articles

The upper section of Figure 5 provides a comprehensive overview of the journals that published systematic reviews citing paper mill articles (i.e., at least one citation). The data show that *International Journal of Molecular Sciences* (*n* = 12), *Cancers* (*n* = 9), and *Medicine* (*n* = 9) were among the academic journals with the highest number of systematic reviews citing paper mill articles. Notably, the number of such publications has increased since 2020.

The right section of Figure 5 illustrates the annual trend of journals publishing these systematic reviews (with at least one citation of paper mill articles). The data indicate a growing trend in Q1 journals, with the number of such systematic reviews increasing from 1 in 2016 to 27 in 2023. Similarly, Q2 journals exhibited a comparable trend, increasing from 0 in 2016 to 18 in 2023. For Q3 journals, the number of publications rose from 2 in 2016 to 12 in 2023.

## 4. Discussion

Systematic reviews play a crucial role in the life sciences by synthesizing high-quality evidence and providing guidance for clinical practice and subsequent research ^33^. Consequently, the literature evaluated and analyzed within systematic reviews must be both accurate and of high quality ^34^. Although numerous studies have expressed concerns about the quality of the literature analyzed in systematic reviews ^35^ ^36^ ^37^ ^38^ ^39^, few have specifically focused on the quality of the evidence these reviews incorporate. With the occurrence of large-scale retraction events triggered by paper mills ^40^, the systematic fabrication of these papers has drawn significant attention from the academic community ^41^ ^42^. However, few studies have examined whether paper mill products have impacted systematic reviews, which inherently require high-quality evidence. By using the references cited in systematic reviews as a point of departure, we conducted the first comprehensive investigation of this issue and provided novel evidence.

We performed a large-scale analysis of the references cited by 100,000 systematic reviews in the life sciences. These references were matched against articles retracted due to paper mills, as recorded in the Retraction Watch Database. Our findings indicate that, although systematic reviews in the life sciences have not been extensively contaminated by paper mill articles (contamination rate: 0.179%), there is a trend of progressively increasing contamination. The number of contaminated systematic reviews rose from four in 2016 to 58 in 2023. This trend parallels the escalating number of retractions linked to paper mills ^43^. The contamination of systematic reviews by paper mill articles may be in an early, “pre-epidemic” stage, necessitating collaborative efforts by the academic community to curb this upward trend.

Our further analysis revealed that the retraction of paper mill articles does not mark the end of their lifecycle. We identified 61 instances where paper mill articles continued to be cited in systematic reviews even after retraction, suggesting that authors of these reviews did not thoroughly verify the retraction status of cited articles. This finding aligns with previous reports ^44^. Another, more challenging issue is that some citations occurred before the retraction, and these systematic reviews did not issue corrections after the cited paper mill articles were retracted. This lack of correction may be partly related to inadequate or nonstandard retraction notices ^45^.

Our analysis of research fields suggests a pattern of concentration in the contamination of systematic reviews by paper mill articles. Oncology, molecular biology, general & internal medicine, cell biology, pharmacology & pharmacy, and environmental sciences & ecology are research areas more prone to contamination. Previous studies have identified oncology and molecular biology as heavily affected areas with respect to retractions ^46^. Our findings further expand on this conclusion, revealing that, in addition to these two fields, multiple other disciplines may also be experiencing contamination. However, continued long-term monitoring and observation will be necessary to confirm this trend.

We also examined systematic reviews that were heavily contaminated (i.e., those citing three or more paper mill articles). Visualization revealed that these reviews might share a similar citation pattern, characterized by extensive clustering of references in the results section. Among these contaminated systematic reviews, the four with the highest number of paper mill article citations were published in journals under controversial academic publishers. Previous research has shown that journals under such publishers exhibit excessively high self-citation rates, potentially signaling problematic citation practices ^47^. Our study provides further evidence that the systematic reviews they publish may deviate from best practices in referencing and evidence synthesis, demonstrating a lack of adequate consideration regarding evidence quality. Additionally, we found that these four systematic reviews had each been cited more than 10 times, with one even cited in government regulations related to the life sciences. This suggests that such systematic reviews could influence subsequent research and policy-making, warranting increased vigilance from the academic community.

Of particular concern is that 76.67% of the systematic reviews citing paper mill articles two or more times were published in high-impact academic journals (Q1 and Q2). A previous study ^18^ investigating 1,182 retracted articles due to paper mills found that 44.8% of them were published in Q2 journals. Our study also identified a similar pattern, reflecting that high-impact academic journals may need to undertake more comprehensive evaluations of systematic reviews, especially scrutinizing the quality of the evidence these reviews synthesize.

Considering the pursuit of perfect evidence quality in systematic reviews and their importance in the life sciences, we believe it is necessary to issue corrections and retractions for systematic reviews currently citing paper mill articles. In particular, for those citing only a single paper mill article, publishing a corrected version after revising the content may be feasible. For reviews heavily contaminated by paper mill articles, academic journals may need to re-assess their quality to ensure the reliability of their conclusions.

We recommend that academic journals, when reviewing systematic articles, match their cited references against databases recording retracted articles, such as Retraction Watch Database, or employ software like “scite” ^48^ to scan for citations of articles retracted due to paper mills. In doing so, retracted articles would not continue to be included as high-quality evidence in systematic reviews post-retraction. Unlike articles retracted for other reasons—such as authorship disputes, conflicts of interest, lack of ethical approval, plagiarism, or partial data fabrication and falsification—articles retracted due to paper mills often contain entirely fabricated and falsified conclusions ^49^ ^50^. Such articles pose a potentially devastating threat to the quality of evidence in systematic reviews. The academic community must take collective action to contain the spread of this contamination in systematic reviews.

Our cross-sectional study has several limitations. We could only match the collected systematic reviews with paper mill articles already identified and retracted, but a significant number of undiscovered paper mill articles may remain. Thus, the articles we matched likely represent only the tip of the iceberg and may underestimate the contamination rate of systematic reviews in the life sciences. Second, we included only systematic reviews indexed in the SCI-Expanded database of the WoS. Due to WoS’s stringent inclusion criteria, it indexes fewer academic papers than some other databases ^51^, which could also lead to an underestimation of the contamination rate. Future studies that incorporate more databases and other types of literature, such as meta-analyses, could more comprehensively assess the impact of paper mills on life sciences research.

Despite these limitations, our study has several notable strengths. We conducted a comprehensive examination of 100,000 systematic reviews in the life sciences and evaluated whether they were contaminated by paper mill articles. This large-scale assessment is the first of its kind in this field. Our study addresses the fifth key issue identified by the United2Act working group ^20^, namely “how are paper mills affecting science and scholarship”. By providing deeper insights and abundant evidence, our research contributes to efforts in research integrity aimed at addressing the paper mill problem.

## 5. Conclusion

Our study highlights the threat posed by paper mill articles to systematic reviews in the life sciences. Although the current scope of contamination is not extensive, there is a clear trend of increasing spread. The academic community should remain vigilant and take action to ensure that systematic reviews— considered the “gold standard” for evidence synthesis—maintain their quality. Citing paper mill articles not only undermines the credibility of the systematic reviews themselves but may also harm future academic research and clinical practice.

## Data Availability

All data produced in the present study are available upon reasonable request to the authors

