## Supplementary Material 1 for "Citation Contamination of Systematic Review Literature in the Life Sciences by Paper Mills"

import re
import pandas as pd

wos_file = "D:/ "
retracted_file = "D:/ "

def parse_wos_data(file_path):
 wos_articles = []
 current_article = {}
 with open(file_path, 'r', encoding='utf-8') as file:
 for line in file:
 line = line.strip()

 if line.startswith("PT "):
 if current_article:
 wos_articles.append(current_article)
 current_article = {}

 elif line.startswith("TI "):
 current_article["Title"] = line[3:].strip()

 elif line.startswith("AU "):
 current_article["Authors"] = line[3:].strip()

 elif line.startswith("SO "):
 current_article["Journal"] = line[3:].strip()

 elif line.startswith("CR "):
 current_article["References"] = []
 while True:
 line = file.readline().strip()
 if line.startswith("NR") or not line:
 break
 dois = re.findall(r'DOI\s([^\s,;]+)', line)
 current_article["References"].extend(dois)

 elif line.startswith("PY "):
 current_article["PublicationYear"] = line[3:].strip()

 elif line.startswith("PD "):
 current_article["PublicationDate"] = line[3:].strip()

 elif line.startswith("SC "):
 current_article["Subject"] = line[3:].strip()

 if current_article:
 wos_articles.append(current_article)

 return pd.DataFrame(wos_articles)


try:
 print(f"load WOS documents：{wos_file}")
 wos_data = parse_wos_data(wos_file)
except FileNotFoundError as e:
 print(f"can’t find the documents, check：{wos_file}")
 raise e


if "References" in wos_data.columns:
 print("example of")
 print(wos_data["References"].head())
else:
 print("fail to abstract")


wos_data["References"] = wos_data["References"].apply(lambda x: x if isinstance(x, list) else [])
wos_data["PublicationYear"] = wos_data.get("PublicationYear", "Unknown").fillna("Unknown")
wos_data["PublicationDate"] = wos_data.get("PublicationDate", "Unknown").fillna("Unknown")
wos_data["Subject"] = wos_data.get("Subject", "Unknown").fillna("Unknown")


try:
 print(f"load data of RW：{retracted_file}")
 retracted_data = pd.read_csv(retracted_file)
 print(f"success")
 print(f"RW includes {len(retracted_data)}。")
except FileNotFoundError as e:
 print(f" can’t find the documents, check：{retracted_file}")
 raise e


if {"DOI", "OriginalPaperDate", "RetractionDate"}.issubset(retracted_data.columns):
 retracted_data["DOI"] = retracted_data["DOI"].str.strip().str.lower()
 print("example of DOI：")
 print(retracted_data[["DOI", "OriginalPaperDate", "RetractionDate"]].head())
else:
 raise ValueError("can’t find the row（DOI, OriginalPaperDate, RetractionDate），please check the content。")


wos_dois = [doi.lower() for references in wos_data["References"] if references for doi in references]
print(f"WOS DOI：{len(wos_dois)}")
print(f"RW DOI：{len(retracted_data['DOI'].unique())}")


matched_dois = set(wos_dois).intersection(set(retracted_data["DOI"]))
print(f"Matched DOI：{len(matched_dois)}")
if matched_dois:
 print("Example of matched DOI：", list(matched_dois)[:10])


matched_records = []
for index, row in wos_data.iterrows():
 if isinstance(row.get("References"), list):
 references = [doi.lower() for doi in row["References"]]
 matches = set(references).intersection(set(retracted_data["DOI"]))
 for match in matches:
 retraction_info = retracted_data.loc[retracted_data["DOI"] == match]
 for _, ret_row in retraction_info.iterrows():
 matched_records.append({
 "WOS Title": row.get("Title", ""),
 "WOS Authors": row.get("Authors", ""),
 "WOS Journal": row.get("Journal", ""),
 "PublicationYear": row.get("PublicationYear", "Unknown"),
 "PublicationDate": row.get("PublicationDate", "Unknown"),
 "Subject": row.get("Subject", "Unknown"),
 "Matched DOI": match,
 "Original Paper Date": ret_row["OriginalPaperDate"],
 "Retraction Date": ret_row["RetractionDate"],
 })

matched_df = pd.DataFrame(matched_records)


if not matched_df.empty:
 print("Matched results：")
 print(matched_df.head())
 output_file = "D:/ "
 matched_df.to_csv(output_file, index=False)
 print(f"Matched results saved as：{output_file}")
else:
 print("Can’t find any matched results。")
